# What we do in the shadows: Methodologists use a range of synthesis methods when meta-analysis of all results is not possible but describe challenges in planning and selecting methods

**DOI:** 10.64898/2026.07.15.26358140

**Authors:** Miranda S. Cumpston, Sue E. Brennan, Rebecca Ryan, James Thomas, Joanne E. McKenzie

## Abstract

**Introduction:** Systematic review authors commonly encounter situations where the data required for meta-analysis are incompletely reported (e.g. when effect estimates are reported without a measure of precision). In this circumstance, many systematic review authors use a method other than meta-analysis (e.g. vote counting), but rarely describe those methods or the rationale for selecting them. We aimed to investigate what methods authors consider when meta-analysis of all study results is not possible, and what factors influence their decisions.

**Methods:** We interviewed 12 experienced systematic review authors, editors and methodologists, presenting four scenarios in which it was not possible to combine all results using meta-analysis. Scenarios varied in the number and size of included studies, available data, and risk of bias. Participants discussed the methods they considered to summarise, synthesise and present the results; whether they would synthesise available results; and how they would draw overall conclusions.

**Results:** Factors that informed decisions included participants’ overall purpose in conducting synthesis, existing beliefs about study results and synthesis methods, trust in the available data, and the decision-making needs of end users. Participants differed in which synthesis methods to use, whether they would use multiple synthesis methods, and which studies they would analyse with each method.

**Conclusions:** We identified several synthesis methods considered when meta-analysis of all results is not possible, and factors that influence the selection of methods, neither of which are routinely reported. More complete reporting of these methods and the factors informing decisions would allow readers to better understand the decisions made.

**Highlights:** *What is already known:* Systematic review authors commonly encounter situations where the data required for meta-analysis are incompletely reported (e.g. when effect estimates are reported without a measure of precision). Many systematic review authors use a method other than meta-analysis (e.g. vote counting), but rarely describe those methods or the rationale for selecting them.

*What is new:* Interviews revealed that systematic review authors consider a range of methods when meta-analysis of all results is not possible, and multiple factors influence the choice of methods. These methods and the decision-making process are not routinely reported in published reviews.

*Potential impact for Research Synthesis Methods readers:* Authors of systematic reviews should report synthesis methods and the rationale for their methods decisions. This would enable readers to understand the decisions made and support interpretation, appraisal and reproduction of the synthesis.

## 1. Introduction

Systematic reviews are a method intended to synthesise the available evidence to answer a research question, with the aim of informing decision making by clinicians, consumers and policy makers.^1^ Where possible, a core analytical component of systematic reviews is the statistical synthesis of results, which often underpins review conclusions. The most commonly applied synthesis method is meta-analysis of effect estimates,^2–4^ however, it has been estimated that meta-analysis is not used in 35% - 56% of systematic reviews,^2–4^ and a larger percentage of reviews do not use meta-analysis for every outcome.^4^

There are several reasons why meta-analysis may not be used (e.g. concerns about clinical or methodological diversity, concerns about bias in the evidence, or too few studies found). In some circumstances, however, even when such concerns do not apply, it may not be possible to include results for all the primary studies in a meta-analysis; for example, when the data required for meta-analysis (usually an effect estimate and measure of precision) may be incompletely or not reported, and cannot be calculated from the available data. Alternatively, different studies may report effect measures for an outcome that are incompatible with each other (e.g. odds ratios and hazard ratios).^3,5–7^ In such situations, alternative statistical synthesis methods have been proposed (here defined as any statistical method used to synthesise numerical results), including summary statistics (e.g. median, range), combining P values, and vote counting based on the direction of effect. These methods can be augmented by tables and visual displays of data (e.g. harvest plots, albatross plots). In addition, structured summaries of the results of individual studies and their characteristics (e.g. risk of bias assessments) can be informative alongside statistical synthesis, or when no synthesis is used.^6,7^

Studies investigating synthesis methods used in systematic reviews have found that beyond meta-analysis, methods are generally not specified or described.^3,4^ Yet, in the absence of meta-analysis, review authors still draw conclusions across study results.^4^ In such circumstances, the methods used to draw conclusions and the underlying rationale for choosing one method ahead of another remain unknown to the reader. Gaining an understanding of the range of synthesis methods considered and applied by review authors, and thereby how the conclusions in published reviews are drawn, is required. Furthermore, understanding the factors that influence decision-making about when to use meta-analysis, other synthesis methods, or a combination of methods may be helpful for informing future guidance on the selection and use, as well as complete and accurate reporting of these methods.

In this study, we aimed to identify:

- the methods for synthesis and presentation of quantitative data considered by experienced review authors when meta-analysis is not possible for some or all studies; and
- the rationale for, and factors that influence, these methods choices.

## 2. Methods

### 2.1. Rationale for chosen study design

We interviewed a sample of experienced systematic review methodologists and authors. We chose interviews as our methodology because other statistical synthesis methods are often incompletely described or absent from systematic reviews.^3,4^ Furthermore, interviews provide the opportunity to investigate authors’ reasoning for their choices, which are also rarely reported.^8,9^ Ethics approval was granted by the Monash University Human Research Ethics Committee (no. 29145). This study is reported in accordance with the COREQ checklist for reporting qualitative studies.^10^

### 2.2. Eligibility and recruitment

Participants were identified through their publication record as authors of systematic reviews of health interventions or exposures and methodological research on systematic reviews, and their participation as members of international professional networks for methodologists such as Cochrane Methods Groups (https://methods.cochrane.org/methods-groups), the GRADE Working Group (www.gradeworkinggroup.org/) and the Campbell Collaboration (www.campbellcollaboration.org/). We aimed to include participants with expertise across a range of systematic review methods (e.g. statistical analysis, systematic reviews of complex interventions) and content areas (e.g. clinical, public health, social interventions and environmental health).

Individuals were invited to participate via email, using publicly available contact information. Individuals wishing to participate in the study responded by email to indicate their consent and schedule the interview. Individuals who did not respond received one reminder email after two weeks. We aimed to include a minimum of 10 participants, up to a maximum of 20 participants or until no new methods were identified.

### 2.3. Development of scenarios for discussion

We developed four scenarios to prompt discussion during the interviews. These were based on a hypothetical systematic review question: ‘*What are the effects of pedometers compared to no intervention on body mass index (BMI)?*’. Each scenario presented a set of studies and their available data, which varied in completeness of reporting (see example scenario in Table 1, and all scenarios in Supplementary File 1, Section A).

**Table 1.** Example of an interview scenario (Scenario 1).

| No. | Study name | N | Available data for outcome BMI |
| --- | --- | --- | --- |
| 1 | Aittasalo 2019 | 181 | Reduced mean BMI by 0.6 kg/m<sup>2</sup> (SE = 1.11) compared with control |
| 2 | Carr 2016 | 40 | MD -1.50 (95% CI -4.49 to 1.49) |
| 3 | Forbes 2018 | 300 | Reduced mean BMI by 10% in the intervention group and 5% in the control group, P = 0.042 |
| 4 | Linde 2022 | 1405 | No effect on BMI (P = 0.142) |
| 5 | Maruyama 2017 | 62 | 50% of participants reduced their BMI by at least 5% in the intervention group, compared with 16% in the control group, P = 0.004 |
| 6 | Morgan 2021 | 110 | Reduced BMI but the result was not statistically significant (at the 5% level) |

Across the four scenarios, we varied several factors to investigate whether they influenced participants’ decisions about synthesis methods (see Table 2 for differences between scenarios). Variations included the number and size of the studies, and the number of studies and participants for which complete data were available for inclusion in meta-analysis. Scenarios 1 to 3 progressively added complexity, through the addition of studies and other information (including risk of bias and the age of participants in each study). The focus of Scenario 4 differed in that we sought to explore participants’ approaches to summarising and presenting the results when statistical synthesis was not appropriate (due to most studies being at a high risk of bias).

**Table 2.**
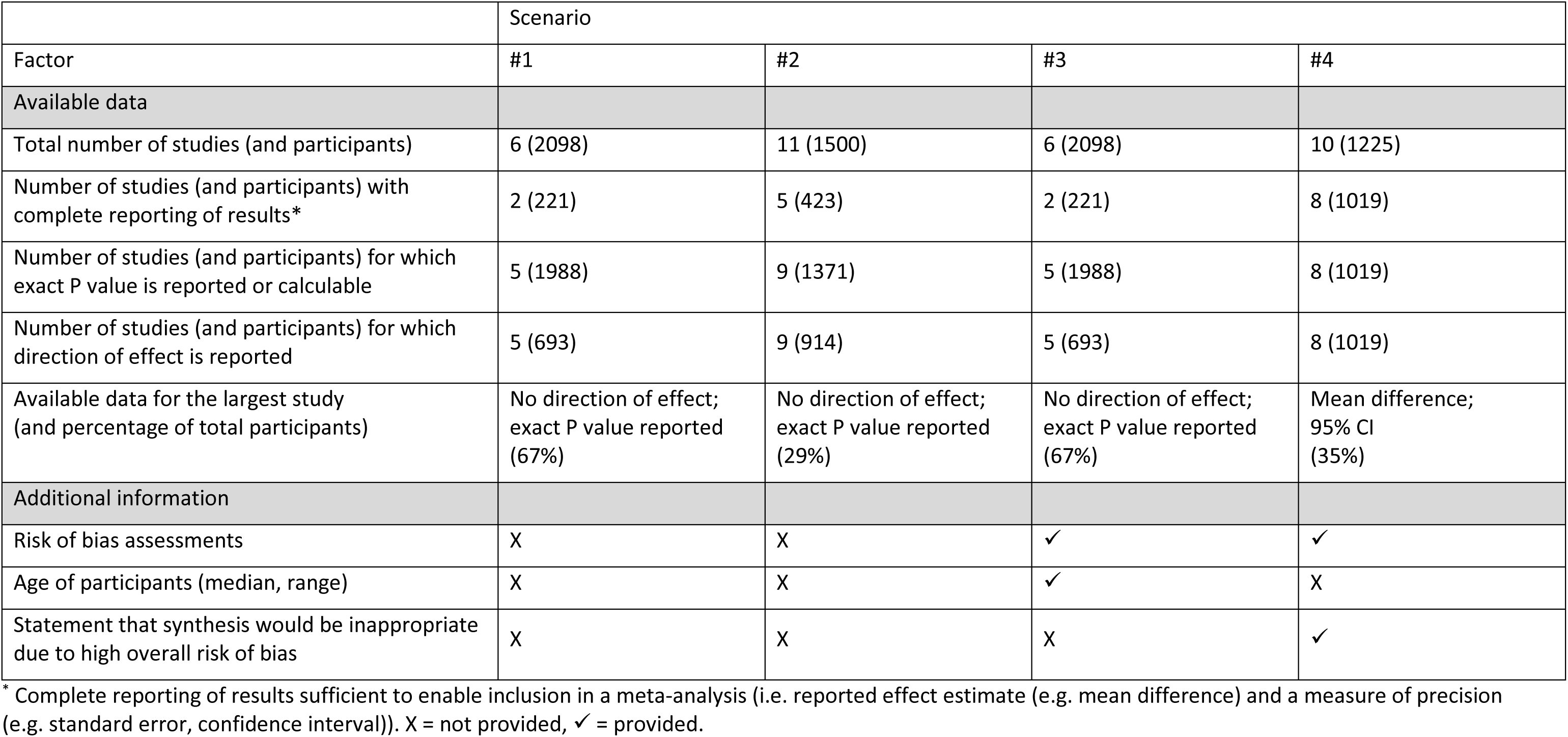
Varying factors between data scenarios presented in the interviews.

|  | Scenario |  |  |  |
| --- | --- | --- | --- | --- |
| Factor | #1 | #2 | #3 | #4 |
| Available data |  |  |  |  |
| Total number of studies (and participants) | 6 (2098) | 11 (1500) | 6 (2098) | 10 (1225) |
| Number of studies (and participants) with complete reporting of results* | 2 (221) | 5 (423) | 2 (221) | 8 (1019) |
| Number of studies (and participants) for which exact P value is reported or calculable | 5 (1988) | 9 (1371) | 5 (1988) | 8 (1019) |
| Number of studies (and participants) for which direction of effect is reported | 5 (693) | 9 (914) | 5 (693) | 8 (1019) |
| Available data for the largest study (and percentage of total participants) | No direction of effect; exact P value reported (67%) | No direction of effect; exact P value reported (29%) | No direction of effect; exact P value reported (67%) | Mean difference; 95% CI (35%) |
| Additional information |  |  |  |  |
| Risk of bias assessments | X | X | ✓ | ✓ |
| Age of participants (median, range) | X | X | ✓ | X |
| Statement that synthesis would be inappropriate due to high overall risk of bias | X | X | X | ✓ |

\* Complete reporting of results sufficient to enable inclusion in a meta-analysis (i.e. reported effect estimate (e.g. mean difference) and a measure of precision (e.g. standard error, confidence interval)). X = not provided, ✓ = provided.

We piloted the scenarios and interview script by interviewing one author (RR), who was not involved in designing the scenarios. The scenarios were slightly amended over the course of the interviews to improve the focus of the scenarios on the availability of data rather than other study characteristics (e.g. ensuring a clear causal link between the intervention and outcome, and removing older publication dates), and to improve the logical order of concepts discussed in the interviews (see Supplementary File 1, Section B).^9^

### 2.4. Interview structure

Prior to interview, participants were sent a copy of the scenarios with a description of the interview process and questions to be discussed (see Supplementary File 1, Section A). As the scenarios were relatively complex, participants were encouraged to read the scenarios ahead of time to consider their responses.

Interviews were conducted via videoconference by two or three authors (MC, JM and for most SB), and lasted a maximum of one hour. A script was used (by MC) to summarise the project and present each scenario. Participants were invited to ‘think aloud’ about the data presented and the methods they would consider to summarise, synthesise and present the results, including:

- how they would interpret the result of each individual study;
- whether they would combine all the available studies in a synthesis;
- how they would combine the study results in a synthesis;
- how they would draw an overall conclusion for the outcome; and
- how they would present the synthesis in a review.

Participants were encouraged to describe the reasoning behind each choice, including any methods they considered but would not use.

After the interview, participants were sent a link to access the scenarios online via a secure platform (Qualtrics^11^) to submit additional comments if they wished.

### 2.5. Research team

All members of the research team had experience undertaking systematic reviews, researching methods, and authoring guidance on systematic review methods,^12,13^ including research and guidance on synthesis methods other than meta-analysis.^14,15^ For this reason, the researchers were in most cases known to the interview participants ahead of the interview: in some cases having collaborated together professionally, and in other cases by reputation. Participants were reassured, in the study documentation and interview, that there is currently no consensus on the best methods for any given scenario, there are no ‘correct’ answers, and the interview was not a test of their knowledge.

Members of the team have experience in collecting and analysing qualitative data from interviews in studies of research methodology (SB, JM) or other qualitative research (SB, MC, RR, JT).

### 2.6. Data and analysis

Interviews were recorded and notes were taken by SB or MC. Notes were checked against the recording by MC to generate a detailed but not verbatim transcript, which was used for analysis. Interview notes and transcripts were not returned to participants.

Informed by guidance for analysis of qualitative data,^16,17^ each interview transcript was reviewed and methods were identified and coded, including methods considered for the summary, synthesis and presentation of results; whether the method considered would or would not be used; factors influencing the selection of methods; and conclusions drawn about the results of individual studies or across studies. Coding was piloted independently for one interview by two authors (MC, JM), and subsequently conducted by MC. For each interview, concepts were initially coded separately for each scenario, and then combined across all four scenarios. Any issues of uncertainty arising during coding were discussed and agreed among three authors (MC, JM, SB). Concepts were then combined across all interviews into a conceptual summary. A figure based on this summary was then prepared, reviewed by all authors and revised accordingly.

## 3. Results

We invited 18 participants: one did not respond, five declined to participate, and 12 accepted our invitation and were interviewed (between July and September 2022). Participants included statisticians, epidemiologists, editors and guideline experts, all with expertise in systematic review methodology. Their fields of research included clinical care, public health, social interventions and environmental health. Following these interviews, we concluded that conceptual saturation had been reached and no new insights about methods were likely to emerge.

We present a visual summary of our analysis of the interviews, structured by steps in the synthesis process (from planning to reporting) in Figure 1. Within each of these steps, we present a summary of the methods elements considered by the interviewees (blue boxes). The summary also incorporates factors influencing decisions: some that would apply irrespective of the review, and some specific to the review (orange boxes left and right, respectively). The figure only includes the range of concepts identified in the interviews and therefore it should not be assumed that the figure provides a comprehensive list of methods, or that the identified methods are endorsed. In the following sections, we highlight key methods elements considered by participants, and identify the factors that influenced the synthesis decisions.

**Figure 1.**
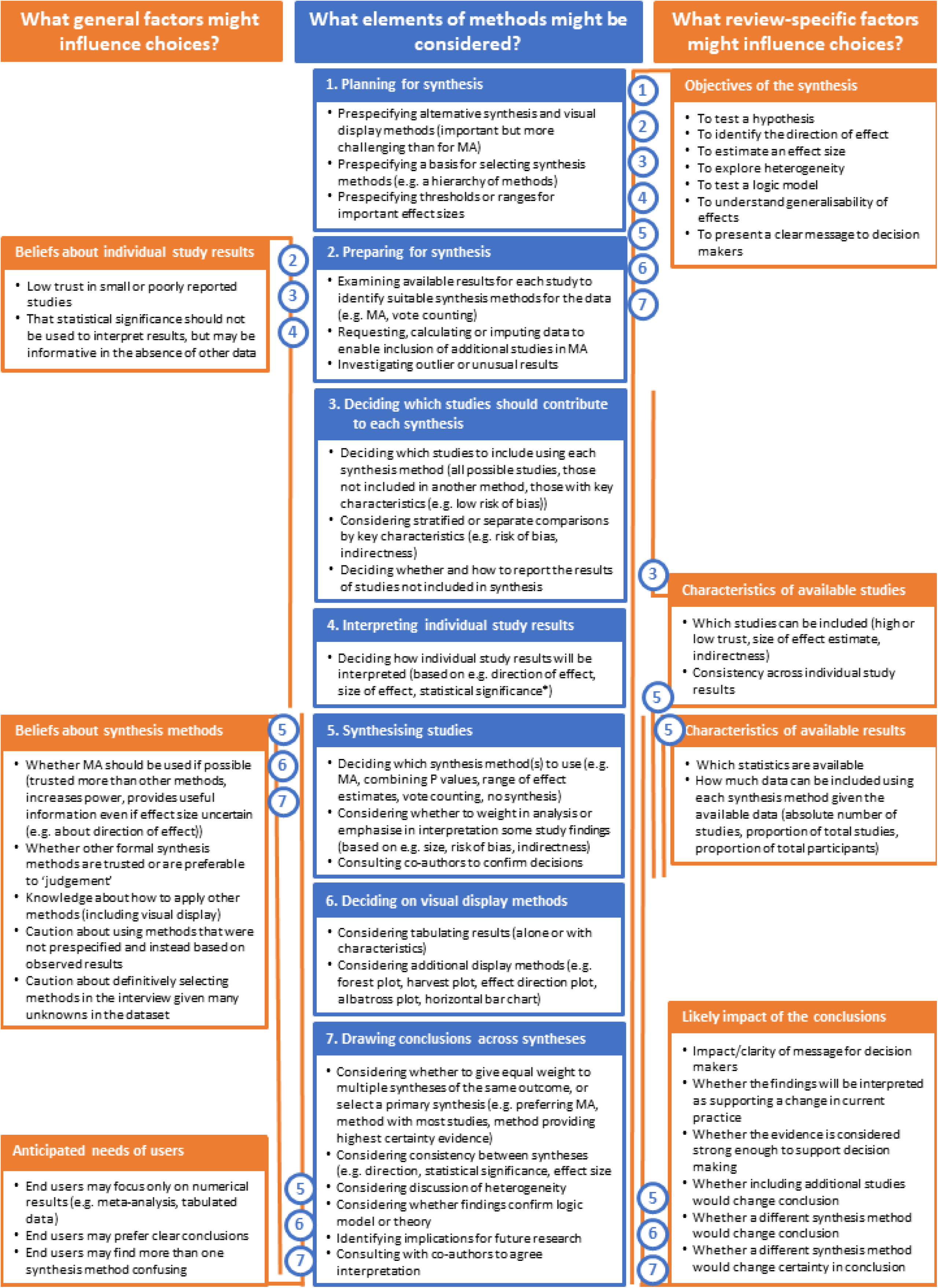
Summary of methods considered and influencing factors as identified by interview participants when meta-analysis of all included studies was not possible. Orange lines and blue numbered circles (corresponding to boxes in the centre column) indicate which steps in the synthesis process are influenced by the factors listed. MID = minimally important difference. PICO = population, intervention, comparator, outcome, methodological characteristics. * Vote counting based on statistical significance is not an acceptable method of synthesis. Cumpston et al. What we do in the shadows: Methodologists use a range of synthesis methods when meta-analysis of all results is not possible but describe challenges in planning and selecting methods.

### 3.1. Planning for synthesis

Some participants noted that planning and prespecification of methods at the protocol stage is an important principle for systematic reviews.

> *“I’m a big fan of consistency, if you have a set of rules, you apply them… [not] making the rules up as you stare at data.”*

> *“Maybe we should be prespecifying more.”*

However, they acknowledged that it is not always possible to anticipate the methods that will be required in a given review.

> *“Some of it I may have prespecified, but I think a lot of it, it’s not until you come to see the data that you see what the challenges are going to be.”*

Some participants suggested that available synthesis methods other than meta-analysis and a process for selecting methods could be prespecified, for example as a hierarchy of alternative methods, anticipating the possibility that meta-analysis may not be possible for all outcomes or results. However, most reported that they rarely or never did this themselves.

> *“Where that’s [meta-analysis] not possible we will use these other approaches [other synthesis methods], make sure there’s some kind of hierarchy in the data analysis plan”.*

> *“Every protocol that I’ve ever put on paper for a systematic review of effects has always focused on our intention for meta-analysis…. I’ve never actually gone that next step… in the event that meta-analysis is impossible or cannot be performed.”*

Some indicated that they prespecified the use of ‘narrative’ synthesis (or similar terminology) where meta-analysis could not be used, which does not provide details of what this entails or how conclusions will be drawn across study results.

> *“I’d be thinking about this from my protocol. Anticipate I can’t do meta-analysis, but I might be able to make sense of the data, so certain features of populations, exposures, outcomes… that I would specify in advance that I would use for organising my qualitative analysis.”*

### 3.2. Preparing for synthesis

Interview participants were asked to assume that the data presented for each study was all that was available, and that the studies had sufficiently similar PICO components (population, intervention, comparison, outcome) and methodological characteristics to combine in the same synthesis.

However, some participants reported that as a first step, they would do their utmost to obtain or calculate the required data for a meta-analysis (e.g. by requesting data from the authors of primary studies, calculating effect estimates and standard errors from the available results, or imputing missing statistics).

> *“Have we exhausted every possibility of transforming data or getting data from the author?*

> *…. My instinct would be to deep dive into metrics used and any transformations possible.”*

Furthermore, participants reported a need to ensure that the primary studies were sufficiently comparable in terms of the PICO components for synthesis.

> *“I would want to know who was in each intervention, the type of intervention, duration in terms of weeks and days, when these measures were taken in order to determine what I would do with the data. Without that information I wouldn’t meta-analyse this information because I wouldn’t know if the interventions are comparable.”*

### 3.3. Selecting studies for synthesis

Some participants noted the importance of reporting all available results in some manner in the review, although it may not be possible to include them all in a single synthesis. Others noted that in some circumstances results not included in meta-analysis may be excluded from any synthesis.

> *“We do have to deal with the other five studies [not included in the meta-analysis]. Wouldn’t want to throw them out…. These are the studies we’ve got, they say something about the direction of effect, we need to incorporate that in some way.”*

> *“All the [reviews] I’m involved in tend to be really big… If we can’t get an effect size, there’ll be a line in the paper that says, we couldn’t get an effect size and we threw them [the additional studies] away.”*

Where participants adopted both meta-analysis and a second synthesis method, some reported that they would include all possible results in each synthesis (meaning some studies would appear in both), while others would limit the second synthesis to the results that could not be included in meta-analysis.

> *“I would include all of the studies [in another synthesis method] even though I have done a meta-analysis. What we would want to see is the whole body of evidence, wherever possible, being compared to each other for the fullest picture possible.”*

> *“I would have one set of studies that I’d used [in] a conventional meta-analysis, and one set of studies that didn’t go into that meta-analysis, where I’d look only at direction of effect. I think because I’ve already analysed the information in the meta-analysis.”*

Some participants noted that results considered untrustworthy (e.g. at high or critical risk of bias, or with extremely poor reporting) may be excluded from synthesis, although some reported that they might apply different study inclusion criteria conditional on the chosen synthesis method (e.g. excluding studies at high risk of bias only when using methods other than meta-analysis).

> *“If the studies were so poor, really can’t get anything from them, this would be a table in the back.”*

### 3.4. Interpreting individual study results

Participants differed in the information they used to interpret each study result, considering one or more of the direction of effect, statistical significance, and size of the effect. Some participants interpreted the effect size in relation to a threshold of clinical or patient importance.

> *“Purely around direction.”*

> *“Statistically significant in favour of intervention, versus no effect, versus statistically in favour of control.”*

> *“I would speak to people who knew what they were talking about and [ask] is that a large effect, is that meaningful to clinicians?”*

In interpreting individual study results, some participants noted that statistical significance alone was not sufficient to interpret a result, but some also noted that in the absence of complete reporting, it was difficult to avoid interpreting a non-significant result as an indicator of limited or no effect.

> *“I’m looking at the direction and magnitude of effect, but also confidence intervals to see whether they are consistent with an increase or decrease, and the thing we are not supposed to do but we all do - whether they [the confidence intervals] cross the null or not.”*

> *“I don’t want to count how many are significant versus not. It’s going to be hard [not] to do that.”*

Participants drew on a range of beliefs and experiences that influenced how they interpreted individual study results. Some expressed a lack of trust in the findings of small studies, studies with incomplete results reporting, results they considered implausible or those they deemed to have ‘unusual’ reporting (e.g. studies reporting the proportion of participants achieving a threshold change in the outcome in each group, rather than the mean of the outcome), and would be less concerned if such results were omitted from synthesis.

> *“But there usually is something about the [smaller] studies that makes me nervous about doing too much with them…. It’s not that that information is not useful, it’s just that there are possibly other factors that I would want to take into account….”*

> *“Why not just report the BMI? I don’t like this data [reporting the proportion of participants achieving a threshold change]. It looks like they’ve cherry-picked it. I’d be very concerned about using it.”*

> *“Quite a significant reduction in the control. What is happening to the control group? I’d want to look more carefully. Seeing big effects in control groups strikes me as a contradiction with the risk of bias. That’s quite an important effect for nothing being administered.”*

### 3.5. Deciding on synthesis methods

All participants considered the use of meta-analysis for synthesis in at least some of the scenarios. Other statistical synthesis methods were rarely named, but included vote counting (based on direction of effect, the size of effect in relation to a threshold of clinical importance, or statistical significance), combining P values, range of effect estimates, or no synthesis (reporting the individual study results only). Some participants selected different methods in different scenarios. All participants expressed uncertainty about the most appropriate methods to use.

> *“[It’s] just not as straightforward to know what to do in these circumstances.”*

Some participants indicated that lack of experience in applying synthesis methods other than meta-analysis increased their uncertainty:

> *“I’m not hugely familiar with doing that [combining P values] myself, I’ve never done it or written it up, so that would be a first time doing that for me.”*

Participants reported that the objective of the systematic review, and the needs of the decision makers who might use the review, would inform their selection of methods. For example, some noted that where the aim of the review was to estimate the size of an intervention effect (e.g. to inform an economic evaluation), meta-analysis would be more likely to be used, even where not all results could be included. For other reviews, the aim may be to understand factors that may cause heterogeneity between studies, and a single effect estimate may be a lower priority.

> *“Who is the decision maker and what do you want them to do with the information?”*

> *“Health economists need a quantitative estimate of the treatment effect to feed in. If the product that I’m producing is to go into a health economic model, I would really try everything to produce a decent quantitative estimate. If it was more of a literature summary then I’m less driven to produce a quantitative summary.”*

> *“[Reviews I work on] tend to be about heterogeneity. They’re less about, ‘What’s the effect?’. We’re dealing with interventions that are going to depend on context and who’s giving it and what’s going on.”*

Participants had existing preferences for synthesis methods: some would always use meta-analysis if possible, and trusted its findings more than other methods; other participants did not hold this preference. There was less familiarity with synthesis methods other than meta-analysis, although some participants noted that using a formal method is preferable to inform conclusions when meta-analysis cannot be used.

> *“I feel just so much more comfortable with a meta-analysis approach…. one of the key drivers is the objectivity, we are not imposing our beliefs or interpretations on the data. As soon as we step away from being able to do that, I would be concerned that I would look at a plot and my interpretation would be different from another person.”*

Some participants expressed the view that vote counting should not be used (on statistical significance or on any basis).

> *“But that method is wrong, and it’s been shown to be in a lot of methods research…. [vote counting] based on the significant positives and significant negatives, because those are just the ones that are statistically powered for that particular outcome in that particular subgroup.”*

> *“It might not be as cut and dried as vote counting,which I think Ijust did, which you’re not supposed to.”*

In practice, when explaining how they might describe study results, participants often used language consistent with vote counting, even if they had not named vote counting as a method they would use.

> *“I would say of the 10 studies, 6 showed a reduction.”*

Some participants used terms such as ‘narrative’ or ‘qualitative’ synthesis as terminology for describing the results of studies when not using meta-analysis. Some suggested structuring their description using theory or a logic model.

> *“I’d be narratively discussing in the text what this information means.”*

> *“A meta-analysis of the studies that have effect sizes and then a narrative discussion of the others.”*

Participants considered the amount of information available in deciding which synthesis methods to use, including: the absolute number of studies (e.g. in response to Scenario 2, where only two studies could be included in a meta-analysis), the proportion of the total number of studies, or the proportion of the total number of study participants that could be analysed using a particular method. In addition, they reported that their level of trust in the specific studies that could be analysed using a given method would influence the choice of synthesis method.

> *“We do facetiously say that you can do meta-analysis with two [studies], but I’m not entirely sure that knowing the mean of those two studies is helpful. Three studies, would I do the meta-analysis? Probably. I’m of two minds here…. We’ve done this whole systematic review, we have six studies, I don’t know that the mean of two studies is a good representation of the whole.”*

> *“I can’t see [the] point of putting two of them into a meta-analysis. I always think you want three, really, just in terms of getting a variance in your meta-analysis apart from anything else. Especially as you’re leaving out most of the data. Not sure how informative it would be, only having one third of the studies… especially if you’ve got bigger studies that are not included at all if you’re doing that.”*

> *“People have a lot of faith in forest plots and pooled results. I prefer to save those for something that you have a bit more confidence in.”*

Some participants were influenced by the complexity of interpreting both results in cases where more than one synthesis method is used.

> *“If you just use one method it’s easier to describe and easier to write up.”*

> *“Would be tempted still to do a vote count and not a meta-analysis unless I can be sure that I would be able to fairly represent the data alongside each other, and come up with a summary statement that would transparently reflect the contribution of the meta-analysis and the contribution of the non-meta-analysis data. That’s challenging for everyone in every area where this happens.”*

Where more than one method was used, some participants anticipated that readers may prioritise the meta-analysis to inform their interpretation of the review findings, which may not be appropriate.

> *“People give more faith in meta-analysis, don’t always give as much credence to anything that sits alongside that. Something that has to be done quite carefully. I worry about getting the balance right.”*

> *“Dependinghow clear that it is in my mind and how clear I can make it to the reader,whether that meta-analysis stays in my review or not, or gets pushedout to supplementary informationfor anyone who’s interested, I’d need to think about that.”*

Paired with the anticipated needs of users, some participants would consider whether using a synthesis method or including additional studies would change the conclusion of the review.

> *“Pooling the data wouldn’t add anything to your conclusions, so why would you do it?”*

> *“Do I do three methods? Do I do two methods? My bottom line is, what am I telling people*

> *about the information, this data we’ve got, what’s the story? Does it help? Does it improve, [does] people’s BMI get reduced, or does everyone’s BMI on average, in general not change? I don’t know how much advantage that confers to people out there.”*

> *“There may be a slight reduction [in the outcome], but we’re probably not sure because of inconsistency. So probably in same position without the meta-analysis than having done it.”*

### 3.6. Deciding on tabular and visual display methods

All participants reported that they would tabulate individual study results, often alongside other key characteristics such as the risk of bias, sample size, or key PICO characteristics. Various other visual display methods were mentioned, most commonly forest plots, but also harvest plots, effect direction plots, albatross plots and horizontal bar charts. In common with the selection of synthesis methods, some participants indicated that a lack of experience in using alternative display options would limit their selection.

Again, the likely interpretation by readers informed participants’ choice of methods. While some would use forest plots to present the findings of individual studies where meta-analysis was not used, a participant noted that in such circumstances readers may use the forest plots to estimate a (possibly misleading) meta-analysed result.

> *“If you switch the diamond [meta-analytic effect estimate] off or not, people are still going to look at the weight of the evidence, is it on this side of the null, that side of the null? The reader is going to synthesise it and make their own judgement.”*

### 3.7. Drawing conclusions across syntheses

Participants found drawing overall conclusions challenging when they used more than one synthesis method, or one synthesis accompanied by individual study results. Some participants considered the findings from the two syntheses equally, while others identified a primary synthesis to form the basis of conclusions.

> *“You could calculate the effect sizes for about half of the studies, so when you’re discussing and writing up your findings, you’d give equal space, or equal weighting to the MA and the other findings.”*

> *“Where possible, we will attempt to assess whether our findings are consistent with the meta-analysis. Implicitly there we’re using the MA as the benchmark, because that is the aspect of the results where we have used a recognised synthesis methodology.”*

When assessing the consistency of results across synthesis methods, participants compared, for example, the direction of effect or statistical significance of each synthesis. Some participants indicated they would consult with co-authors to reach appropriate conclusions.

> *“You’d want to have more than one or two people overall [discussing the findings], to come to a decision you’re comfortable with.”*

## 4. Discussion

### 4.1. Key findings

Our interview study of 12 experienced systematic review authors and methodologists identified that when meta-analysis of all results is not possible, considerable uncertainty exists. Participants were uncertain about which synthesis method(s) to use, whether they would use multiple synthesis methods or opt for one, and what studies they would include using different methods. Furthermore, participants found it challenging to draw conclusions across more than one synthesis result, or a synthesis result alongside individual study results. Participants described a range of factors influencing their selection of synthesis methods and the interpretation of results, including beliefs and experience, characteristics of the dataset, and communication to decision makers. While some participants articulated rules or heuristics for selecting particular methods that could be pre-specified (e.g. never meta-analysing only two studies, or presenting a hierarchy of preferred synthesis methods), others felt that they would need to see the available data to decide which synthesis methods would be most appropriate.

Our findings demonstrate that even when experienced systematic reviewers are presented with the same dataset, their approach to synthesizing and interpreting the findings was sometimes very different, potentially leading to different conclusions. Lorenc *et al* observed that researchers have to negotiate a difficult pathway when selecting research methods in circumstances where no single method may be demonstrated to be appropriate or clearly superior to others.^18^

### 4.2. What this study adds to what is already known

The practices of researchers in the field of evidence synthesis have been explored using interviews in other studies.^18–20^ Of most relevance to our study was that of Lorenc *et al*, in which the authors aimed to understand the factors that may influence researchers’ selection of synthesis methods in the presence of complexity (e.g. in the aims, methods, interventions and contexts of the studies in a review). Similarly to our findings, they found that decisions about methods were made largely in an informal way, drawing on the participants’ own expertise and judgement, and further noted that these decisions may not be declared in systematic reviews.^18^

Methods studies focusing on synthesis methods other than meta-analysis used in systematic reviews have found that the use of these methods is common (occurring in more than half of published reviews for one or more outcomes), but that specific methods of synthesising results are rarely described.^3,4^ Authors of published reviews may use broad terms such as ‘narrative synthesis’ to describe their methods, but in practice the most common method observed for drawing conclusions across studies was vote counting, whether or not this method is named in the review.^4^ This is consistent with the present study’s finding that while a range of synthesis methods are considered, details of the chosen approach are often not reported.

### 4.3. Strengths and limitations

The study benefitted from experienced participants, who work across multiple disciplines and areas of health research. The interview method, incorporating specific scenarios and ‘think aloud’ methods, enabled us to investigate and identify ‘hidden’ methods that are considered and incorporated into synthesis, but rarely reported in published reviews.^18^ Although our sample was small, we were able to reach saturation of new ideas.

The interviews in this study were limited to considering a small number of scenarios, each of which included a limited number of studies, for feasibility of discussion. Larger datasets with different characteristics may have elicited different responses. In comparison to a real review, our participants lacked in-depth knowledge of the included studies and the time to consider their decisions and consult with colleagues.

It is possible that methods and approaches expressed by participants were influenced by social desirability bias. However, participants were experienced researchers and openly conveyed their uncertainties, suggesting that social desirability bias may have had a limited effect.

### 4.4. Implications of this research

Authors of systematic reviews should plan at protocol stage for the possibility that meta-analysis cannot proceed for some or all of the included studies in the review. A hierarchy of preferred methods and a process for selecting methods for the review could be specified, acknowledging that different methods may be appropriate in different circumstances, and no universally applicable hierarchy of the ‘best’ methods is available. Both prespecified and post hoc decisions can be described in the methods of completed reviews. More complete reporting of methods would allow readers to understand and critique the selected synthesis methods, and provide models for other authors encountering similar scenarios.

Participants identified a knowledge gap and lack of confidence in selecting and applying other synthesis methods and visual displays that is likely to be widespread, given the level of experience of the participants in this study. Additional training and guidance resources may assist in resolving this gap, including worked examples, computer code to implement the methods, and guidance on what to report, may be of value.

## 5. Conclusion

We identified synthesis methods used when meta-analysis of all results is not possible, and factors that influence the selection of methods, neither of which are routinely reported in published reviews. Influencing factors identified included beliefs and experience, characteristics of the dataset, and the needs of decision makers. More complete reporting of synthesis methods and the factors informing decisions would be possible, allowing readers to better understand the decisions made and the rationale for the decisions.

## Data Availability

The data that support the findings of this study are not publicly available, in accordance with the ethics approval of the study and consent provided by research participants.

## Acknowledgements

With permission, we acknowledge the participants in this study for their time and insights: Edoardo Aromataris, Deborah Caldwell, Davina Ghersi, Theresa Moore, Alison O’Mara-Eves, Terri Piggot, Rebecca Ryan, Nancy Santesso, Natalie Strobel, James Thomas, Hugh Sharma Waddington and Paul Whaley. Rebecca Ryan and James Thomas were part of the team undertaking this research. MC received funding from the Australian Government Research Training Program. JM is supported by a National Health and Medical Research Council Investigator Grant (GNT2009612). SB’s position at Cochrane Australia is funded by the Australian Government through the National Health and Medical Research Council. RR’s position at Cochrane Consumers and Communication is supported by funding from the Australian Government through the National Health and Medical Research Council. Funding organisations had no role in the conduct or reporting of this study. JT is supported in part by the National Institute for Health and Care Research. This report is independent research supported by the National Institute for Health and Care Research ARC North Thames. The views expressed in this publication are those of the author(s) and not necessarily those of the National Institute for Health and Care Research or the Department of Health and Social Care.

## CRediT author statement

Miranda S. Cumpston: Methodology, Formal analysis, Investigation, Data Curation, Writing – Original Draft, Writing – Review & Editing, Project Administration. Sue E. Brennan: Conceptualization, Methodology, Investigation, Validation, Writing – Review & Editing. Rebecca Ryan: Investigation, Validation, Writing – Review & Editing. James Thomas: Investigation, Validation, Writing – Review & Editing. Joanne E. McKenzie: Conceptualization, Methodology, Validation, Formal Analysis, Investigation, Data Curation, Writing – Review & Editing, Supervision.

## Supplementary File 1

## Section A: Interview discussion scenarios (final)

**Summary and synthesis when meta-analysis is not possible for all or some studies**

## Introduction

We are interested in identifying the full range of approaches used by experienced systematic review authors when meta-analysis is not possible for some or all studies. This may include both formal and informal methods and thought processes that may not be described in published papers.

### What will we ask you to do in your interview?

During the interview, we will present you with the scenarios in this document, and ask you to describe your approach, the factors you take into consideration and the reasoning behind your approach. Each scenario includes data from several studies.

We are interested to hear about the following:

- How would you interpret the result of each individual study?
- Would you combine all the available studies in a synthesis?
- How would you combine the study results in a synthesis?
- How would you draw an overall conclusion for this outcome?
- How would you present this synthesis in a review (e.g. text or visual displays)? There is no need to perform any analysis, only to talk about what you would do.

We use the same systematic review question throughout but the studies and available data vary. We are interested in whether the changes to the data lead you to change your approach and, if so, why.

Cumpston et al. What we do in the shadows: Methodologists use a range of synthesis methods when meta-analysis of all results is not possible but describe challenges in planning and selecting methods

### The review question used in all scenarios

*What are the effects of pedometers compared to no intervention for weight loss?*

You have no concerns about combining these studies in a synthesis. Specifically, you should assume:

- You are at the point where you are ready to analyse the data.
- You have determined that the population, interventions, comparators and outcome measurement in all the included studies are similar enough to combine in synthesis.
- All the included studies are randomised trials and the statistical methods used in each study are appropriate for the study design.
- You have no concerns about missing results.
- No additional information is available from the published papers or the authors of the studies, so you are restricted to the data presented in the scenario.

The scenarios presented are commonly encountered by systematic review authors. Additional methods such as the investigation of heterogeneity or methods that may facilitate meta-analysis (e.g. conversion of effect measures) are not the focus of this study.

Cumpston et al. What we do in the shadows: Methodologists use a range of synthesis methods when meta-analysis of all results is not possible but describe challenges in planning and selecting methods

### Scenario 1

In a review of pedometer interventions, you include the outcome BMI (kg/m^2^).

In this scenario six studies measured and reported BMI for our comparison of interest. The results are in the table below. Blue cells indicate there is enough data to include the study in a meta-analysis. There are no concerns about combining any of the 6 studies in a synthesis.

**Considering the information below, talk us through your approach to interpreting and synthesising the data.**

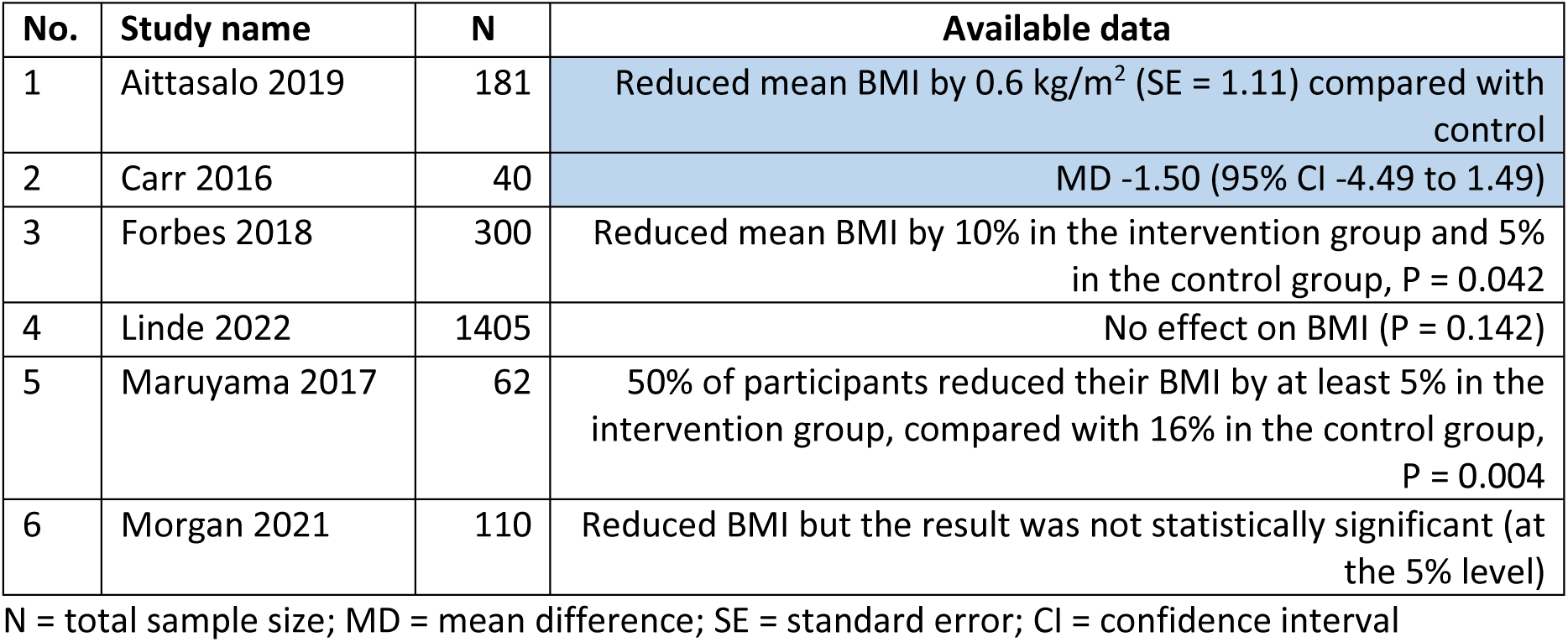

| No. | Study name | N | Available data |
| --- | --- | --- | --- |
| 1 | Aittasalo 2019 | 181 | Reduced mean BMI by 0.6 kg/m<sup>2</sup> (SE = 1.11) compared with control |
| 2 | Carr 2016 | 40 | MD -1.50 (95% CI -4.49 to 1.49) |
| 3 | Forbes 2018 | 300 | Reduced mean BMI by 10% in the intervention group and 5% in the control group, P = 0.042 |
| 4 | Linde 2022 | 1405 | No effect on BMI (P = 0.142) |
| 5 | Maruyama 2017 | 62 | 50% of participants reduced their BMI by at least 5% in the intervention group, compared with 16% in the control group, P = 0.004 |
| 6 | Morgan 2021 | 110 | Reduced BMI but the result was not statistically significant (at the 5% level) |

**When you talk us through your approach, we are interested to know:**

- How would you interpret the result of each study?
- Would you combine all the available studies in a single synthesis?
- How would you combine the study results in a synthesis?
- How would you draw an overall conclusion for this outcome?
- What is the reasoning behind your choices?
- How you would present this synthesis in a review (e.g. text or visual displays)?

Cumpston et al. What we do in the shadows: Methodologists use a range of synthesis methods when meta-analysis of all results is not possible but describe challenges in planning and selecting methods

### Scenario 2

In this scenario, eleven studies measured and reported the outcome of BMI (kg/m^2^) for the comparison of interest. Compared with **Scenario 1**, studies 1 to 6 are the same, except that now the sample size in Linde 2022 is 430, not 1405. Studies 7 to 11 are new. Blue cells in the table indicate there is enough data to include the study in a meta-analysis. There are no concerns about combining any of the 11 studies in a synthesis.

**Considering the information below, talk us through your approach to synthesising the data.**

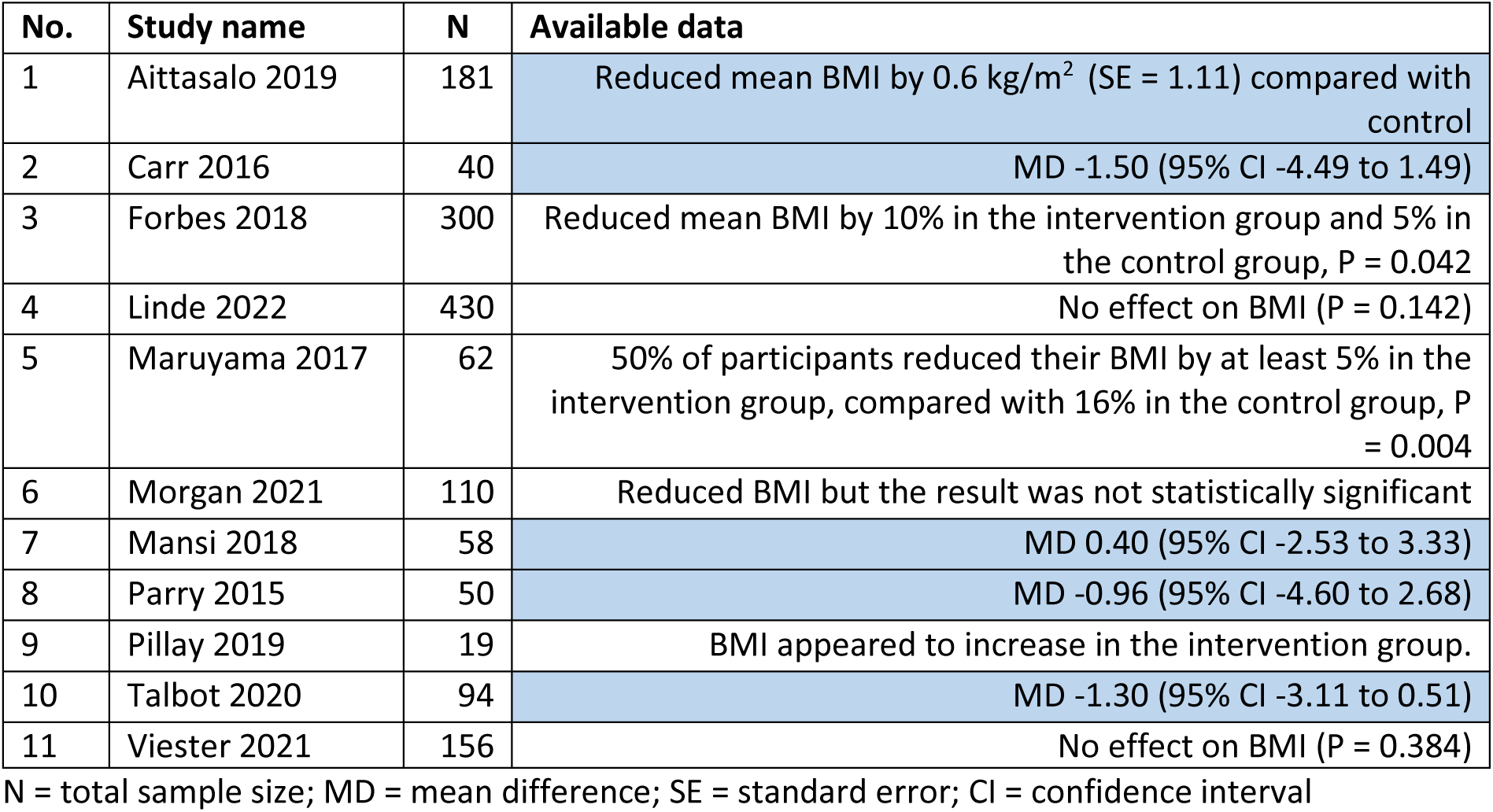

**When you talk us through your approach, we are interested to know**

Compared to **Scenario 1:**

- Would your chosen methods of synthesis change?
- If so, what synthesis methods might you use and why?
- Are there any additional factors you considered? Is there anything different in the reasoning behind your choices?

Cumpston et al. What we do in the shadows: Methodologists use a range of synthesis methods when meta-analysis of all results is not possible but describe challenges in planning and selecting methods

### Scenario 3

In this scenario, the same studies and numerical data are provided as for **Scenario 1**. In addition, information on the overall risk of bias for each study for this outcome and the age of the included participants is provided. Blue cells in the table indicate there is enough data to include the study in a meta-analysis.

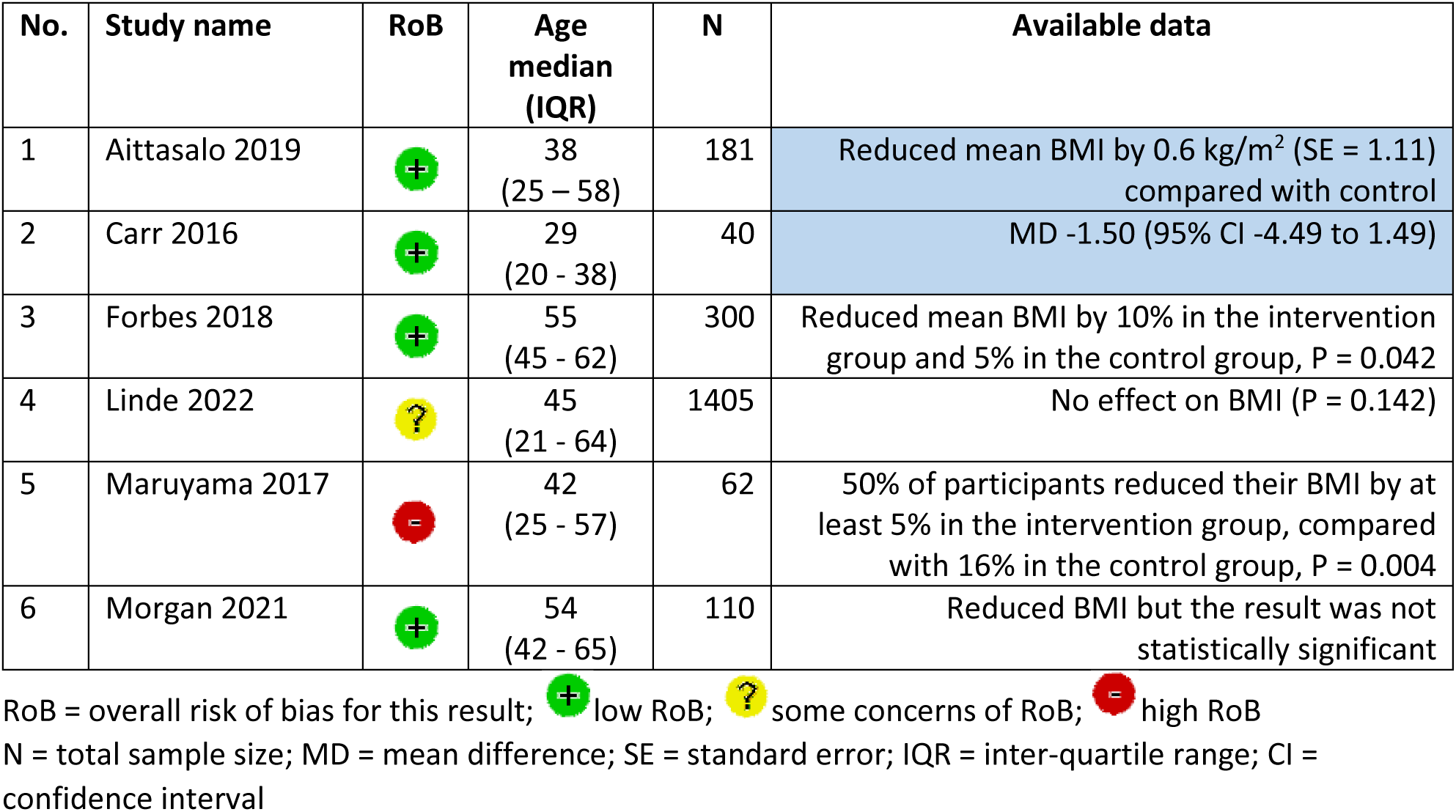

| No. | Study name | RoB | Age median (IQR) | N | Available data |
| --- | --- | --- | --- | --- | --- |
| 1 | Aittasalo 2019 |  | 38<br>(25 – 58) | 181 | Reduced mean BMI by 0.6 kg/m<sup>2</sup> (SE = 1.11) compared with control |
| 2 | Carr 2016 |  | 29<br>(20 - 38) | 40 | MD -1.50 (95% CI -4.49 to 1.49) |
| 3 | Forbes 2018 |  | 55<br>(45 - 62) | 300 | Reduced mean BMI by 10% in the intervention group and 5% in the control group, P = 0.042 |
| 4 | Linde 2022 |  | 45<br>(21 - 64) | 1405 | No effect on BMI (P = 0.142) |
| 5 | Maruyama 2017 |  | 42<br>(25 - 57) | 62 | 50% of participants reduced their BMI by at least 5% in the intervention group, compared with 16% in the control group, P = 0.004 |
| 6 | Morgan 2021 |  | 54<br>(42 - 65) | 110 | Reduced BMI but the result was not statistically significant |

RoB = overall risk of bias for this result; low RoB; some concerns of RoB; high RoB
N = total sample size; MD = mean difference; SE = standard error; IQR = inter-quartile range; CI = confidence interval

Compared to **Scenario 1:**

- Would your chosen methods of synthesis change?
- If so, what synthesis methods might you use and why?

Are there ***any other important considerations*** for synthesising data when meta-analysis is not possible for all or some of the studies that have not been discussed?

Cumpston et al. What we do in the shadows: Methodologists use a range of synthesis methods when meta-analysis of all results is not possible but describe challenges in planning and selecting methods

### Scenario 4

In this scenario, ten studies measured and reported the outcome of BMI (kg/m^2^) for the comparison of interest.

In eight of studies, you have been able to extract (or calculate from available data) a mean difference in BMI between groups (along with a 95% confidence interval). However, in seven of the studies, you judged the result to be at a high risk of bias. Because of this **you conclude that synthesis of this outcome would not be appropriate**.

**Considering the information below, talk us through your approach to summarising the data from individual studies, without synthesis.**

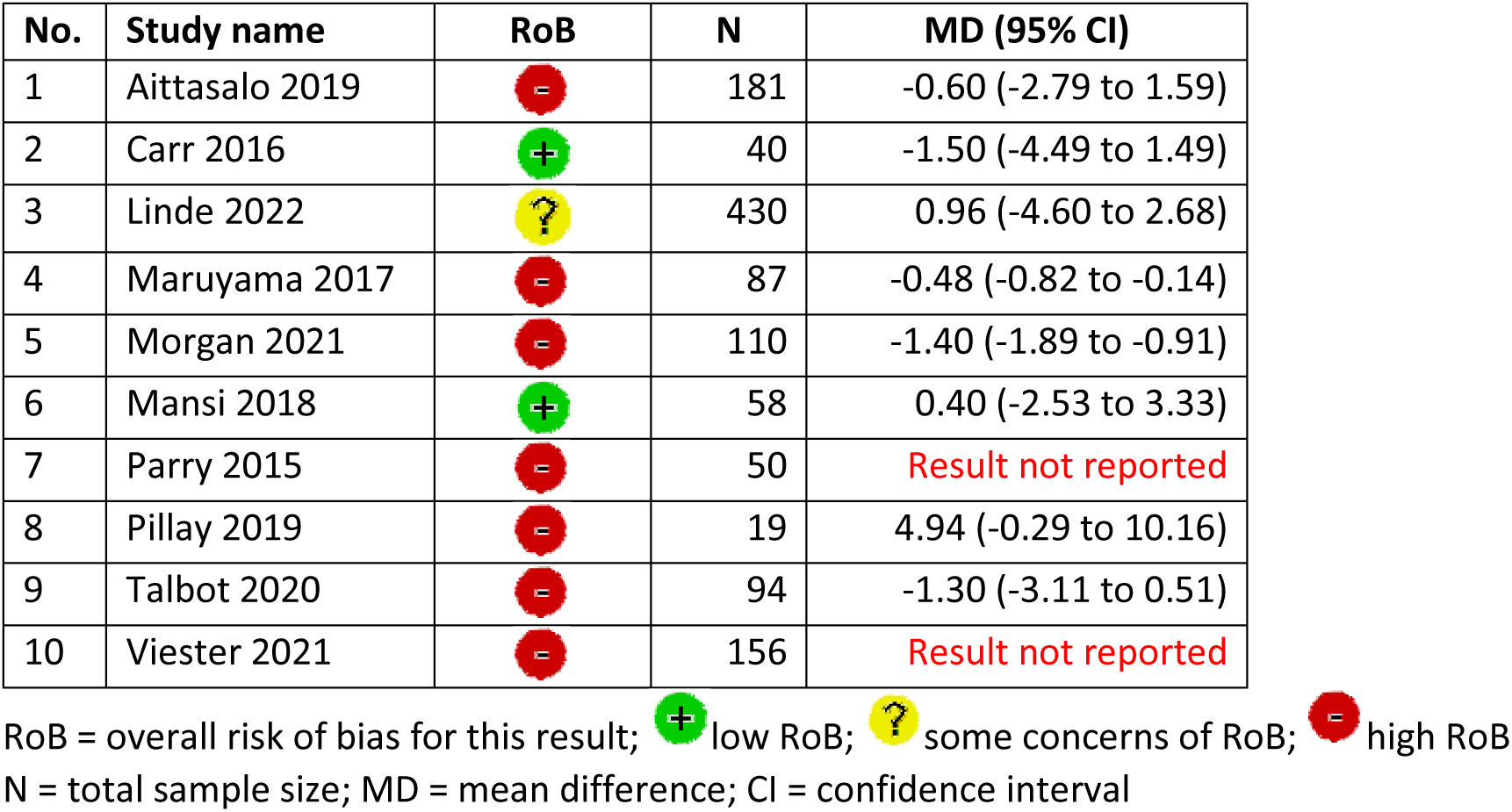

| No. | Study name | RoB | N | MD (95% CI) |
| --- | --- | --- | --- | --- |
| 1 | Aittasalo 2019 |  | 181 | -0.60 (-2.79 to 1.59) |
| 2 | Carr 2016 |  | 40 | -1.50 (-4.49 to 1.49) |
| 3 | Linde 2022 |  | 430 | 0.96 (-4.60 to 2.68) |
| 4 | Maruyama 2017 |  | 87 | -0.48 (-0.82 to -0.14) |
| 5 | Morgan 2021 |  | 110 | -1.40 (-1.89 to -0.91) |
| 6 | Mansi 2018 |  | 58 | 0.40 (-2.53 to 3.33) |
| 7 | Parry 2015 |  | 50 | Result not reported |
| 8 | Pillay 2019 |  | 19 | 4.94 (-0.29 to 10.16) |
| 9 | Talbot 2020 |  | 94 | -1.30 (-3.11 to 0.51) |
| 10 | Viester 2021 |  | 156 | Result not reported |

RoB = overall risk of bias for this result; low RoB; some concerns of RoB; high RoB N = total sample size; MD = mean difference; CI = confidence interval

**When you talk us through your approach, we are interested to know:**

- What method(s) might you use to summarise and present this data?
- Would you draw an overall conclusion for this outcome?
- What factors do you consider? What is the reasoning behind your approach?
- Are there things that you find particularly challenging, or where multiple approaches could be taken?
- How you would present this data in a review (e.g. text or visual displays)?

Cumpston et al. What we do in the shadows: Methodologists use a range of synthesis methods when meta-analysis of all results is not possible but describe challenges in planning and selecting methods

## Section B. Modifications to interview scenarios during study

| Category | Specific change | Rationale |
| --- | --- | --- |
| Changed after pilot interview |  |  |
| Publication year | Publication years in study IDs in each scenario revised to 2015 or later | To avoid speculation on age of study as a reason for differences in reporting and focus discussion on handling the available data. |
| Study order | Order of studies in Scenario 4 revised to match other scenarios. | To increase clarity for participants as to which data were the same across scenarios. |
| Changed after interview 5 |  |  |
| Hypothetical review question | Research question for hypothetical review addressed by all scenarios changed from investigating pedometers for “physical activity” to “weight loss”. | To remove consideration of the directness of BMI as the outcome measure reported in scenarios, and allow focus on handling the available data. |
| Comparability of study characteristics | Text revised to more directly state that the studies included in the scenarios have been determined to be comparable for synthesis based on PICO characteristics, and that there are no concerns about missing data. | To minimise distraction during the interview on speculation about study characteristics and allow focus on handling the available data. |
| Questions for discussion | Revision to questions for consideration to include stepped progression through interpretation of individual studies, decision about which studies to synthesise, selection of synthesis method, drawing overall conclusions and visual display methods. | To increase clarity and logical progression of questions for participants, and ensure each aspect of decision making was explicitly addressed. |
| Order of scenarios | Scenario containing information on risk of bias and age moved to follow the two scenarios without such information (initially at scenario 2, now scenario 3). | To avoid information about risk of bias influencing selection of methods in other scenarios. |
| Sample size | Sample size of largest study (Linde 2022) changed in (new) Scenario 2 and Scenario 4 from 1405 to 430. | To consider selection of methods without any one study being much larger than the others in the scenario. |
| Study characteristics | In Scenario 4, details for largest study (Linde 2022) changed from high to unclear risk of bias, and from no results reported to full results reported. Risk of bias rating and reported results swapped with other studies to retain the same overall risk of bias profile and number of studies not reporting results. | To increase plausibility of scenario, as participants would expect the largest available study to have a lower risk of bias and report results in full. |
| General | Minor revisions to wording throughout. | To increase clarity for participants. |

## Notes

**Funding statement:** MSC received funding for this work from the Australian Government Research Training Program. JEM is supported by a National Health and Medical Research Council Investigator Grant (GNT2009612). SEB’s position at Cochrane Australia is funded by the Australian Government through the National Health and Medical Research Council. RR’s position at Cochrane Consumers and Communication is supported by funding from the Australian Government through the National Health and Medical Research Council. Funding organisations had no role in the conduct or reporting of this study. JT is supported in part by the National Institute for Health and Care Research. This report is independent research supported by the National Institute for Health and Care Research ARC North Thames. The views expressed in this publication are those of the author(s) and not necessarily those of the National Institute for Health and Care Research or the Department of Health and Social Care.

### Competing Interest Statement

The authors have declared no competing interest.

### Author Declarations

The Human Research Ethics Committee of Monash University gave ethical approval for this work.

